# Factors Associated with Anxiety and Depression among Reproductive-aged Women in Bangladesh: Insights from the 2022 DHS

**DOI:** 10.1101/2024.12.30.24319793

**Authors:** Isna Haque Sheoti, Sanzana Saira Shezuti, Md. Zakiul Alam

## Abstract

**Introduction:** Mental health disorder causes significant disturbance to every aspect of an individual’s life. Almost one in every eight people in the world lives with a mental disorder. In this study, we aimed to measure the prevalence and determinants of anxiety and depression among reproductive-aged women in Bangladesh.

**Methodology:** We utilized data from Bangladesh demographic and health survey-2022. Anxiety and depression were measured by the Generalized Anxiety Disorder 7 Scale (GAD-7) and the Patient Health Questionnaire (PHQ-9). We used the chi-square test in bivariate analysis and log-binomial logistic regression to explore the factors associated with anxiety and depression.

**Findings:** Around 20% and 5% had symptoms of anxiety and depression, respectively. Compared to women aged 15-19, anxiety was significantly higher among women ages 20-49, and women aged 45-49 (PR= 1.73, p=0.004) had a substantially higher depression. Women living in Rangpur had the highest prevalence of both anxiety and depression. Women from the poorest households and having a bank account had a significantly higher prevalence of anxiety and depression than others. Muslims had higher anxiety (PR=1.40, p<0.001) and depression (PR= 1.48, p=<0.001). Women with positive attitudes towards wife-beating had higher anxiety (PR=1.28 p<0.001) and depression (PR= 1.47, p=<0.001). Women with no education had significantly higher anxiety levels (PR=1.37, p<0.001), whereas those who had access to mass media had a higher depression (PR=1.20, p=0.016).

**Conclusion:** Women of reproductive age face multiple crucial life events that can have an impact on their mental health. In this situation, strategic policy actions need to be provided at both family, community, and provider levels to safeguard the mental health of women in the country. Additionally, specific actions and programs need to drive for women aged 40 or above, women of lower socioeconomic class, women of Rangpur, and women suffering from domestic violence.

## Introduction

Anxiety is a future-oriented mood state associated with preparation for possible, upcoming adverse events.^1^ It can also be described as the central nervous system’s physiological and emotional responses to a vague sense of threat or danger.^2^ On the other hand, depression involves a depressed mood or loss of pleasure or interest in activities for long periods.^3^ It’s a low, sad state in which life seems dark and has overwhelming challenges.^4^ Anxiety is the most common mental illness in the world, affecting 301 million people.^5^ Approximately 280 million people in the world suffer from depression.^3^ An estimated 3.8% of the population experience depression, including 5% of adults (4% among men and 6% among women) and 5.7% of adults older than 60 years.^3^

Depression and anxiety can affect all aspects of life, including relationships with family, friends, and community.^5–7^ The quality of life among depressed adults is more impaired than that of adults with diabetes, hypertension, and chronic lung disease.^8^ Similarly, anxiety has also been associated with productivity, impaired work, family and social functioning, physical disability, and even mortality.^6,8–10^ Many factors may be associated with anxiety, including the ongoing dangerous societal conditions, poverty, and various societal and cultural pressures, which can help create a climate where the development of anxiety disorder becomes likely.^4,11^ The etiology of depression is complex, and much of it is still not well understood, most notably a history of mental illness, poor social support, the experience of instability and violence, hormonal changes during pregnancy and childbirth, and possibly socioeconomic status and poor physical health.^12^

Women are twice as likely to suffer from mental health disorders.^13^ In 2010, the global prevalence of depression was 5.5% and 3.2%, respectively, representing a 1.7-fold greater incidence in women.^14^ Women’s distinct psychological, neurochemical, anatomic, hormonal, genetic, and personality factors make them more vulnerable to depression and anxiety than men.^15^ Worldwide, about 10% of pregnant women and 13% of women who have just given birth experience a mental disorder, primarily depression. It is even higher in developing countries, i.e., 15.6% during pregnancy and 19.8% after childbirth.^16^ In Bangladesh, one in five women of reproductive age (15-49) have symptoms of anxiety, and 5% of them have symptoms of depression.^17^ Thus, it is crucial to explore a situation analysis of mental health disorders among women in Bangladesh.

In Bangladesh, many studies on the prevalence and determinants of anxiety and depression were conducted. However, they focused on urban-rural differences, the effect of COVID-19, postpartum mothers, older people, adolescents, construction workers, women living in urban slums, expected mothers and fathers, garment workers, and university students.^18,19,28,29,20–27^ However, none of the studies focused on reproductive-aged (15-49) women. In this regard, our study aimed to measure the prevalence and identify the socioeconomic determinants associated with anxiety and depression among reproductive-aged women in Bangladesh.

### Theoretical framework of the study

We followed the ‘Lack of Control’ theory for the study (**Figure 1)**. The sense of personal control is the cognitive connection between emotional distress and social structural conditions.^30^ The sense of personal control is the feeling that one’s own actions and efforts shape one’s life. The sense of control versus powerlessness is the cognitive embodiment of disadvantage, structural inequality, and objective powerlessness. It includes socioeconomic status and its elements (educational attainment, work, income, gender, age, neighborhood, and race/ethnicity). A low sense of personal control leads to anxiety, depression, and anger.^30–32^ While one continuum of personal control is the learned, generalized belief that one can and does master one’s life, the other end consists of the belief that external forces, such as luck, chance, fate, or powerful others, shape one’s life.^33^ As we explored the factors associated with anxiety and depression among reproductive-aged (15-49) women in Bangladesh, we theorized those components of lack of control theory affected the level of depression and anxiety among women in the country.

**Figure 1:**
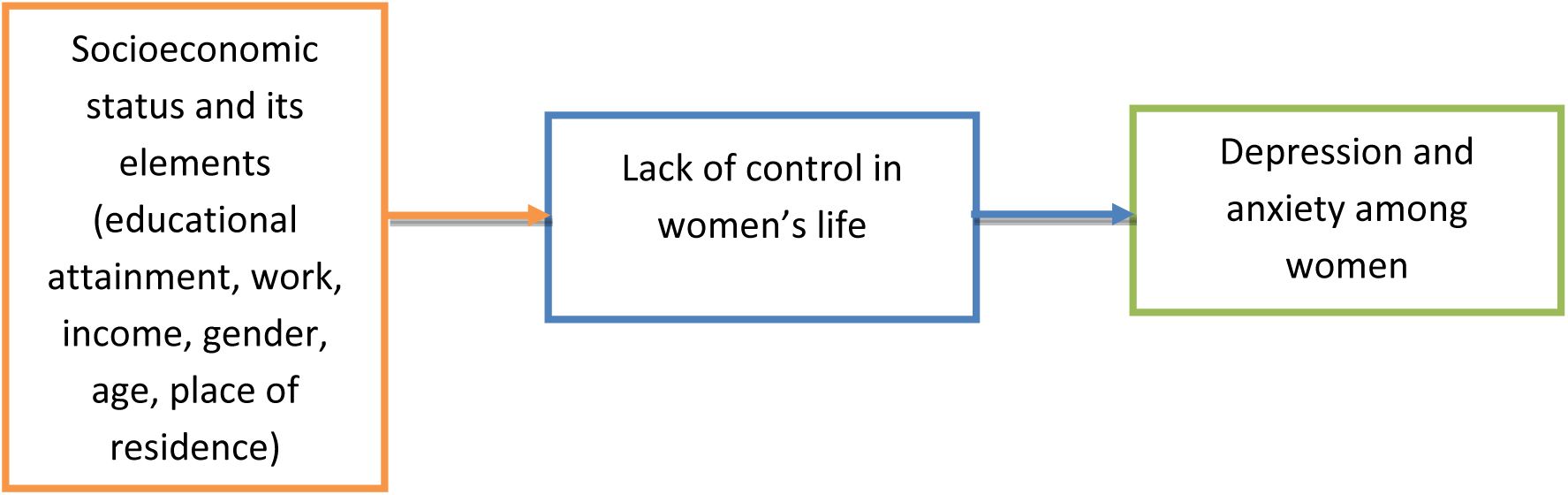
Theoretical framework of this study.

## Data and methods

### Data source

This study used the latest available nationally representative cross-sectional data from the Bangladesh Demographic and Health Survey (BDHS) 2022.^17^ The BDHS sample is nationally representative, and the detailed methodology is elsewhere in the report.^17^ The BDHS follows two-stage stratified sampling with a response rate of 98.6%. **Figure 2** presents the process of sample selection for the study. In the first stage, 30,375 were selected from 675 enumeration areas (EAs). However, the survey collected data from only 674 EAs due to security issues in one EA in Cox’s Bazar. After that, 45 households were selected from each EA, and 30 EAs were selected for the long questionnaire. The analytical sample for this study, was 20,029, which includes mental health and well-being.

**Figure 2:**
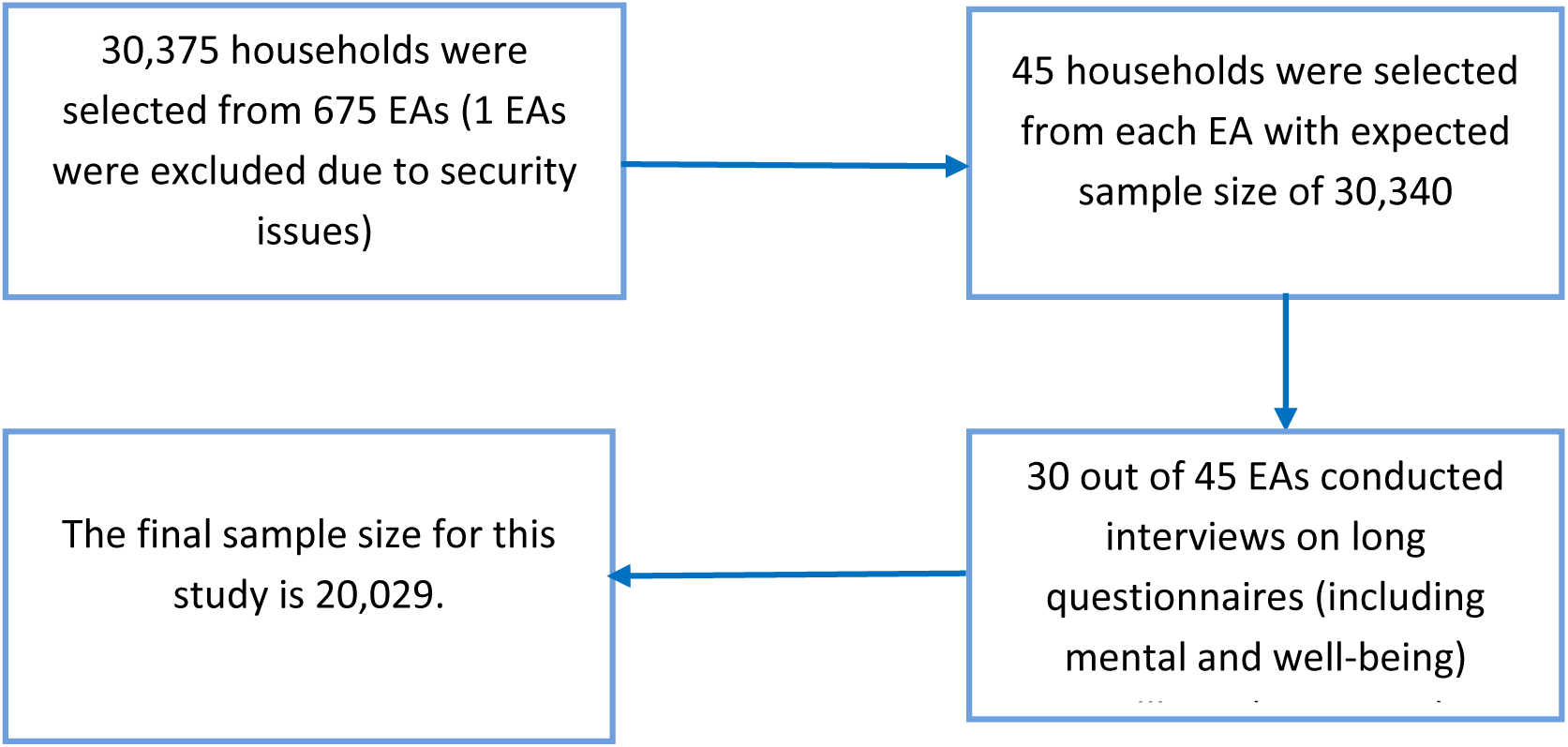
Process of sample selection for the study.

### Variables of study

#### Dependent variables

This study employed two dependent variables: anxiety and depression. BDHS measured anxiety by using the Generalized Anxiety Disorder 7 Scale (GAD-7).^17^ GAD-7 is a set of seven items designed to assess the primary characteristics of anxiety, including persistence and impairing worry.^34^ In addition, the GAD-7 captures attributes related to three other common anxiety disorders, namely, panic disorder, social anxiety disorder, and posttraumatic stress disorder.^34^ The scale focused on symptoms experienced in the 2 weeks preceding the survey **(Figure 3).** The severity of symptoms for anxiety is depicted using a Likert scale in which scores of 0, 1, 2, and 3 are assigned to the response categories ‘not at all’ (never), ‘several days’ (rarely), ‘more than half the days’ (often), and ‘nearly every day’ (always). The total anxiety scale is generated by adding the scores of individual items. The highest score for GAD-7 is 21, and respondents who scored six or higher on the GAD-7 were considered to have symptoms of anxiety.^17,35^ This scale shows an 89% sensitivity and 82% specificity in detecting GAD.^17^

**Figure 3:**
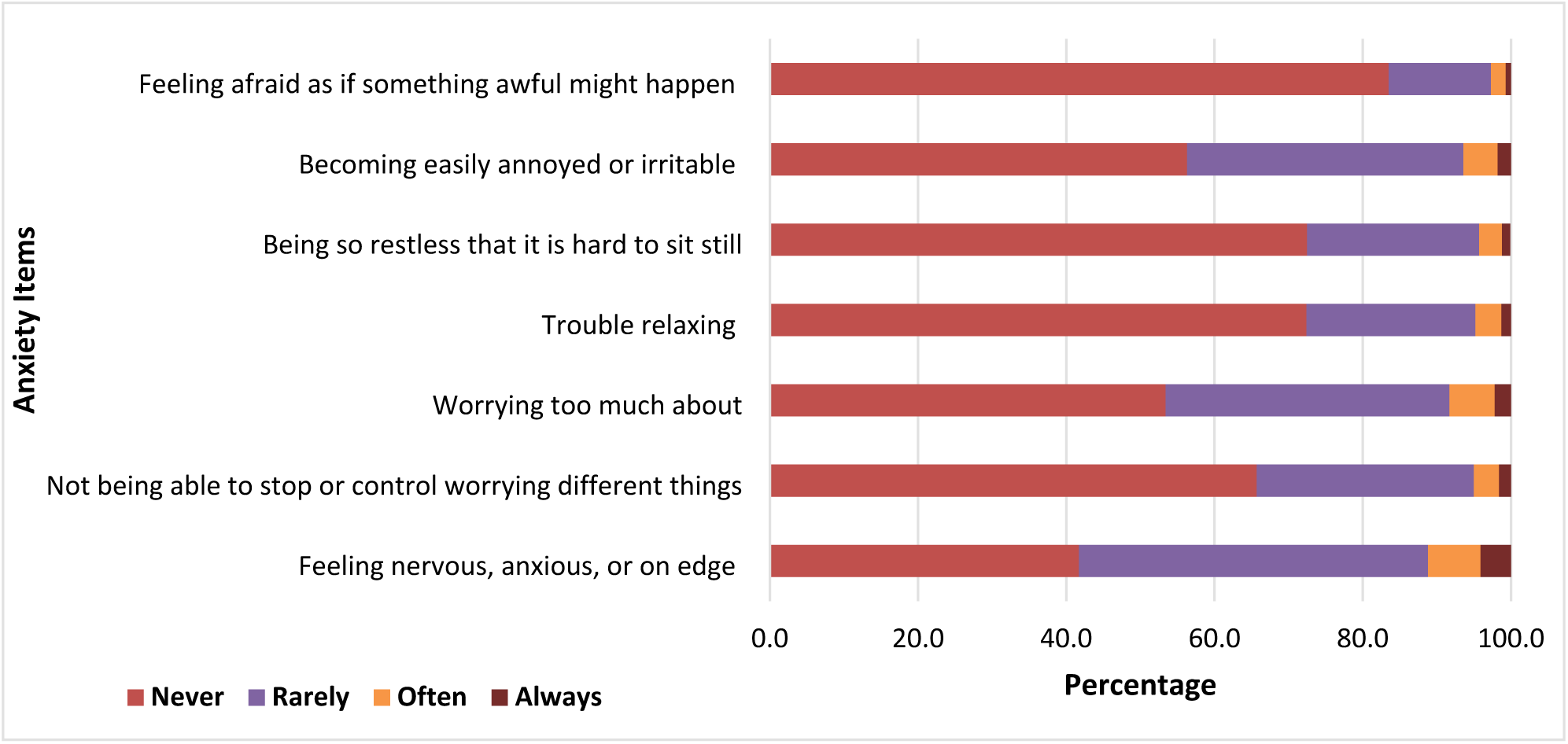
Items used to measure anxiety (%)

Depression was measured using the Patient Health Questionnaire (PHQ-9).^17^ The questions in PHQ-9 are based on the *Diagnostic and Statistical Manual of Mental Disorders (DSM)* criteria for diagnosis of depression.^17,36^ The severity of symptoms for depression is depicted using a Likert scale in which scores of 0, 1, 2, and 3 are assigned to the response categories ‘not at all’(never), several days’ (rarely), more than half the days’ (often), and ‘nearly every day’ (always) **(Figure 4).** The highest score of PHQ-9 is 27, and respondents who scored ten or higher are considered to have symptoms of depression.^17,36^ A score of 10 or more has a sensitivity of 88% and a specificity of 88% for major depression.^17^

**Figure 4:**
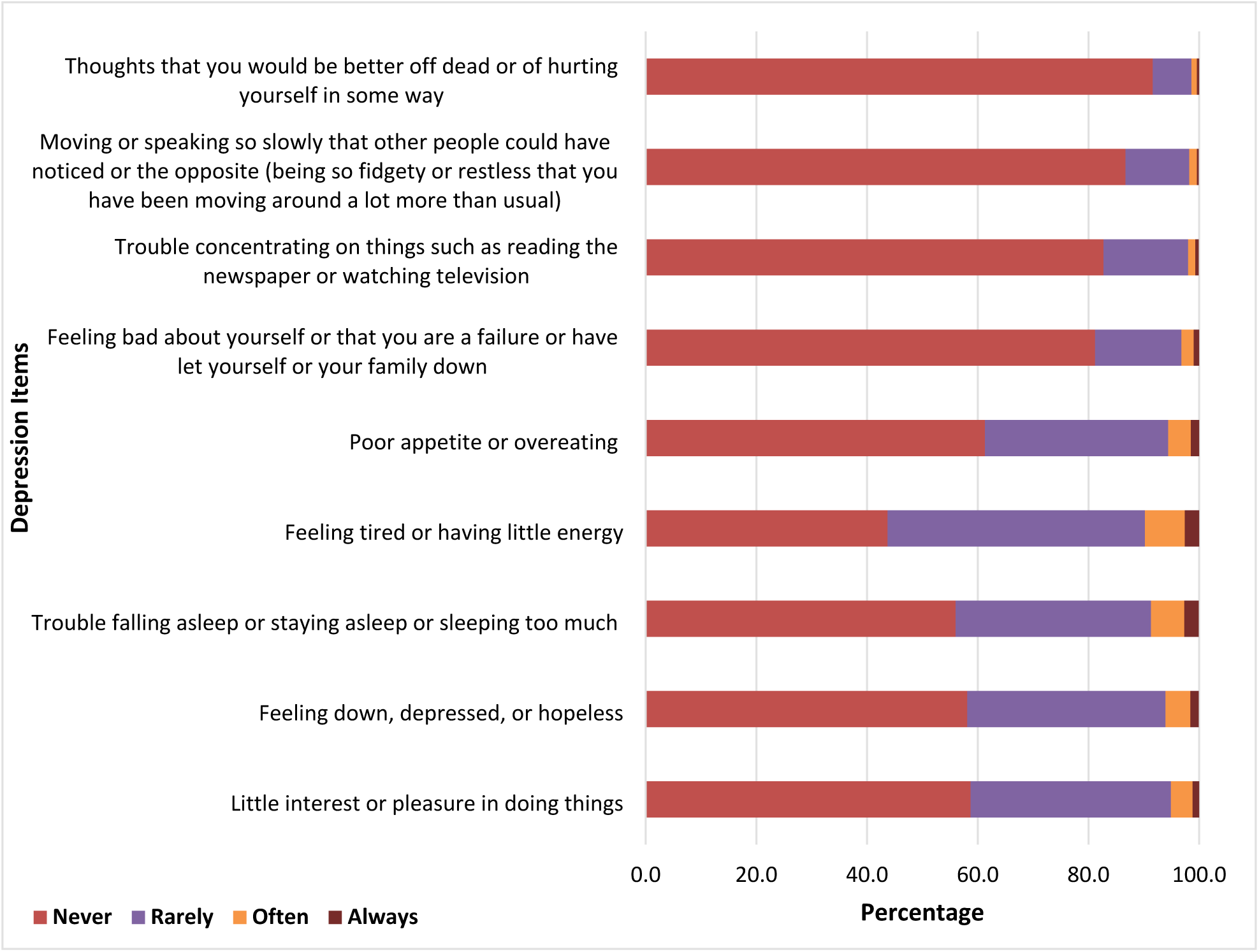
Items used to measure depression (%)

#### Independent variables

The independent variables were selected based on the theoretical model (Figure 1) and the availability of variables in the dataset.^17^ The independent variables are age, division, place of residence, highest educational level, wealth index, working status, having an account in a bank or other financial institution, participation in decision-making, attitudes toward wife beating, husband’s level of education, religion, exposed to any mass media, and ever used internet. We categorized the reproductive-aged women into five years age categories (15-19, 20-24, 25-29, 30-34, 35-39, 40-44, and 45-49). Religion was categorized as Muslim and others (Hindu, Buddhist, or Christian) due to most Muslims in Bangladesh. Administrative division (eight divisions: Barisal, Chattogram, Dhaka, Khulna, Rajshahi, Rangpur, and Sylhet) and place of residence (whether residing in the urban or rural area) are two major spatial factors we included in the study. Education (wife and husband) was categorized as no education, primary, secondary, and higher. The wealth index used to assess the socioeconomic status of the household was constructed from data on household possessions using the principal component analysis and divided into quintiles (poorest, poorer, middle, richer, and richest). The working status of women, having an account in a bank or other financial institution, exposure to any mass media, and ever-used internet were categorized as yes or no.

Two different dimensions of women’s empowerment were used: women’s participation in household decision-making and attitude towards wife-beating justified. We measured women’s participation in household decision-making as a proxy of women’s empowerment using four pieces of information: 1) *the person who usually decides on the respondent’s health care*, 2) *the person who usually determines on large household purchases*, 3) *the person who usually decides on visits to family or relatives*, and 4) *the person who usually decides what to do with money husband earns.* Participation was ‘yes’ if the respondents made all the decisions alone or jointly with the husband or others and ‘no’ if husbands or others made the decision. Attitude toward wife beating justified was measured using five items with the responses of yes, no, and don’t know: *beating justified if* 1) *wife goes out without telling husband,* 2) *wife neglects the children,* 3) *wife argues with husband,* 4) *wife refuses to have sex with husband,* 5) *wife burns the food*. We categorized ‘wife beating justified’ as ‘yes’ (positive) if any responses were ‘yes’ and no (negative) if the responses of all items were ‘no.’

### Statistical analysis

We first analyzed sample characteristics by outcomes (**Table 1**), where p-values were from Rao and Scott’s corrected Chi-square tests.^37^ To explore the factors associated with anxiety and depression, we conducted log-binomial regression. For binary outcomes from cross-sectional data, logistic regression is better fitted and extensively used in literature. When prevalence is more than 5%, the odds ratio (OR) from logistic regression overestimates the prevalence ratio (PR). PR is also preferred for communication. Therefore, we used log-binomial regression. We ran two models [unadjusted and adjusted]. Multicollinearity was checked with variance inflation factor (VIF) using a linear regression. The goodness-of-fit (GOF) was tested using a standard logistic regression model, and the model was found to fit the data. The PR, 95% confidence interval (CI), and associated probability value (p-value) were presented. All the analyses were conducted using Stata 18, considering survey weights and complex design.

**Table 1:** Sample characteristics by anxiety and depression.

| Variables | Anxiety |  | Depression |  | Total |
| --- | --- | --- | --- | --- | --- |
|  | No | Yes | No | Yes |  |
| **N(%)** | 16,126 (80.5%) | 3,902 (19.5%) | 19,042 (95.1%) | 986 (4.9%) | 20,029 (100.0%) |
| **Age in 5-year groups** |  |  |  |  |  |
| 15-19 | 1,506 (9.3%) | 223 (5.7%)* | 1,666 (8.7%) | 63 (6.4%)* | 1,729 (8.6%) |
| 20-24 | 2,866 (17.8%) | 423 (10.8%) | 3,183 (16.7%) | 106 (10.8%) | 3,289 (16.4%) |
| 25-29 | 2,925 (18.1%) | 598 (15.3%) | 3,360 (17.6%) | 163 (16.5%) | 3,523 (17.6%) |
| 30-34 | 2,720 (16.9%) | 716 (18.4%) | 3,259 (17.1%) | 178 (18.0%) | 3,437 (17.2%) |
| 35-39 | 2,559 (15.9%) | 785 (20.1%) | 3,162 (16.6%) | 182 (18.5%) | 3,344 (16.7%) |
| 40-44 | 1,948 (12.1%) | 598 (15.3%) | 2,409 (12.7%) | 137 (13.9%) | 2,546 (12.7%) |
| 45-49 | 1,601 (9.9%) | 559 (14.3%) | 2,002 (10.5%) | 157 (15.9%) | 2,160 (10.8%) |
| **Mean (SD)** | 31.34 (8.9) | 33.92 (8.6)* | 31.74 (8.9) | 33.80 (8.8)* | 31.84 (8.9) |
| **Division** |  |  |  |  |  |
| Barishal | 973 (6.0%) | 226 (5.8%)* | 1,143 (6.0%) | 56 (5.7%)* | 1,199 (6.0%) |
| Chattogram | 2,947 (18.3%) | 802 (20.5%) | 3,592 (18.9%) | 157 (15.9%) | 3,749 (18.7%) |
| Dhaka | 4,237 (26.3%) | 843 (21.6%) | 4,869 (25.6%) | 211 (21.4%) | 5,080 (25.4%) |
| Khulna | 1,909 (11.8%) | 480 (12.3%) | 2,233 (11.7%) | 156 (15.8%) | 2,389 (11.9%) |
| Mymensingh | 1,267 (7.9%) | 260 (6.7%) | 1,470 (7.7%) | 57 (5.8%) | 1,527 (7.6%) |
| Rajshahi | 2,158 (13.4%) | 467 (12.0%) | 2,527 (13.3%) | 98 (10.0%) | 2,625 (13.1%) |
| Rangpur | 1,703 (10.6%) | 587 (15.0%) | 2,117 (11.1%) | 174 (17.7%) | 2,291 (11.4%) |
| Sylhet | 932 (5.8%) | 237 (6.1%) | 1,093 (5.7%) | 75 (7.7%) | 1,169 (5.8%) |
| **Place of residence** |  |  |  |  |  |
| Urban | 4,678 (29.0%) | 1,023 (26.2%)* | 5,449 (28.6%) | 251 (25.4%) | 5,700 (28.5%) |
| Rural | 11,449 (71.0%) | 2,880 (73.8%) | 13,593 (71.4%) | 735 (74.6%) | 14,328 (71.5%) |
| **Highest educational level** |  |  |  |  |  |
| No education | 2,023 (12.5%) | 729 (18.7%)* | 2,564 (13.5%) | 189 (19.1%)* | 2,752 (13.7%) |
| Primary | 4,065 (25.2%) | 1,149 (29.4%) | 4,947 (26.0%) | 267 (27.1%) | 5,214 (26.0%) |
| Secondary | 7,682 (47.6%) | 1,677 (43.0%) | 8,914 (46.8%) | 445 (45.1%) | 9,359 (46.7%) |
| Higher | 2,356 (14.6%) | 347 (8.9%) | 2,617 (13.7%) | 86 (8.7%) | 2,703 (13.5%) |
| **Mean (SD)** | 6.918 (4.197) | 5.886 (4.097)* | 6.757 (4.197) | 5.936 (4.137)* | 6.717 (4.198) |
| **Wealth index** |  |  |  |  |  |
| Poorest | 2,785 (17.3%) | 798 (20.5%)* | 3,381 (17.8%) | 202 (20.5%) | 3,583 (17.9%) |
| Poorer | 3,158 (19.6%) | 869 (22.3%) | 3,821 (20.1%) | 207 (20.9%) | 4,028 (20.1%) |
| Middle | 3,345 (20.7%) | 790 (20.2%) | 3,912 (20.5%) | 223 (22.6%) | 4,135 (20.6%) |
| Richer | 3,399 (21.1%) | 790 (20.2%) | 3,997 (21.0%) | 192 (19.4%) | 4,189 (20.9%) |
| Richest | 3,439 (21.3%) | 654 (16.8%) | 3,931 (20.6%) | 163 (16.5%) | 4,094 (20.4%) |
| **Working status** |  |  |  |  |  |
| No | 11,035 (68.4%) | 2,582 (66.2%)* | 12,957 (68.0%) | 659 (66.8%) | 13,617 (68.0%) |
| Yes | 5,091 (31.6%) | 1,321 (33.8%) | 6,085 (32.0%) | 327 (33.2%) | 6,412 (32.0%) |
| **Has an account in a bank or other financial institution** |  |  |  |  |  |
| No | 12,986 (80.5%) | 3,044 (78.0%)* | 15,292 (80.3%) | 738 (74.8%)* | 16,030 (80.0%) |
| Yes | 3,141 (19.5%) | 858 (22.0%) | 3,750 (19.7%) | 249 (25.2%) | 3,999 (20.0%) |
| **Participation in decision-making** |  |  |  |  |  |
| Low (No) | 4,949 (30.7%) | 1,170 (30.0%) | 5,794 (30.4%) | 325 (32.9%) | 6,119 (30.6%) |
| High (Yes) | 11,177 (69.3%) | 2,732 (70.0%) | 13,248 (69.6%) | 662 (67.1%) | 13,910 (69.4%) |
| **Attitudes toward wife beating** |  |  |  |  |  |
| Yes (Positive) | 2,342 (14.5%) | 773 (19.8%)* | 2,894 (15.2%) | 221 (22.4%)* | 3,115 (15.6%) |
| No (Negative) | 13,785 (85.5%) | 3,129 (80.2%) | 16,148 (84.8%) | 766 (77.6%) | 16,913 (84.4%) |
| **Husband's level of education <sup>a</sup>** |  |  |  |  |  |
| No education | 3,205 (19.9%) | 1,081 (27.7%)* | 4,012 (21.1%) | 274 (27.7%)* | 4,286 (21.4%) |
| Primary | 4,522 (28.0%) | 1,147 (29.4%) | 5,382 (28.3%) | 287 (29.1%) | 5,669 (28.3%) |
| Secondary | 5,363 (33.3%) | 1,176 (30.1%) | 6,233 (32.7%) | 306 (31.0%) | 6,539 (32.6%) |
| Higher | 3,036 (18.8%) | 499 (12.8%) | 3,415 (17.9%) | 120 (12.2%) | 3,535 (17.6%) |
| **Religion** |  |  |  |  |  |
| Others | 1,625 (10.1%) | 296 (7.6%)* | 1,849 (9.7%) | 72 (7.3%) | 1,921 (9.6%) |
| Muslim | 14,501 (89.9%) | 3,606 (92.4%) | 17,193 (90.3%) | 914 (92.7%) | 18,107 (90.4%) |
| **Exposed to any mass media** |  |  |  |  |  |
| No | 6,692 (41.5%) | 1,740 (44.6%)* | 8,022 (42.1%) | 411 (41.6%) | 8,432 (42.1%) |
| Yes | 9,434 (58.5%) | 2,162 (55.4%) | 11,021 (57.9%) | 576 (58.4%) | 11,596 (57.9%) |
| **Ever used internet** |  |  |  |  |  |
| No | 11,047 (68.5%) | 2,952 (75.7%)* | 13,250 (69.6%) | 750 (76.0%)* | 14,000 (69.9%) |
| Yes | 5,079 (31.5%) | 950 (24.3%) | 5,792 (30.4%) | 237 (24.0%) | 6,029 (30.1%) |

Frequency (%): The column percentage is in parentheses except for total, N(%). N(%) is a row percentage. P-value from Rao and Scott corrected Chi-square tests. \*Significant at $\leq 0.05$ level
Statistics were computed using survey weights and complex design.
a. There was some missing data (69) and the "don't know" category (13). It was imputed with the Hot-deck method.

### Ethics Statement

This study is based on de-identified secondary data from BDHS, so it does not require IRB approval. The 2022 BDHS was conducted by the National Institute of Population Research and Training (NIPORT) of the Ministry of Health and Family Welfare, where Mitra and Associates of Bangladesh implemented the survey. ICF International of Calverton, Maryland, USA, provided technical assistance. Data was collected with prior permission, where an interview was conducted only if the respondent provided their verbal consent in response to being read out an informed consent statement by the interviewer. The NIPORT took ethical approval from the Bangladesh Medical Research Council (BMRC) for the survey. The data set is available at https://dhsprogram.com/data.

## Results

The sample characteristics of the participants are presented in **Table 1**. The highest percentage of respondents was 25-29 (17.6%), and the lowest was 15-19 (8.6%). Most of the respondents were from rural areas (71.5%). Almost half of the respondents completed secondary education (46.7%), and 32.6% of respondents’ husbands had secondary-level education. The lowest respondents were from the poorest social class (17.9%). While 68% of respondents did not work, 32% worked. Nearly all (80%) did not have an account in a bank or other financial institution. While 30.6% of women did not participate in decision-making (low participation), 15.6% justified wife-beating (negative attitude). Most of the respondents were Muslim (90.4%). While 57.9% of respondents had access to mass media, access to the internet was low (30.1%).

**Table 2** presents factors associated with anxiety among participants using log-binomial regression. Women from all age groups had significant differences with women aged 15-19 in their prevalence of anxiety. Compared to women aged 15-19, the prevalence of anxiety was significantly higher among those aged 25-29 [PR:1.32, p<0.001], 30-34 [PR:1.62, p<0.001], 35-39 [PR:1.82, p<0.001], 40-44 [PR:1.82, p<0.001], and 45-49 (PR=1.77, p<0.001). Women living in Rangpur had the highest prevalence (PR=1.57, p<0.001) and significantly differed from Mymensingh. Women with no education significantly differed in their anxiety level (PR=1.37, p<0.001). Women who belonged to the poorest (PR=1.21, p=0.012) and poorer (PR=1.21, p=0.006) economic classes had a significantly higher prevalence. Women who had an account in a bank or other financial institution (PR=1.25, p<0.001) had a considerably higher prevalence than those who didn’t. Women with a positive attitude towards wife beating had 28% higher levels of anxiety (PR=1.28, p<0.001) than women with a negative attitude. Muslim (PR=1.40, p<0.001) respondents had a higher prevalence of anxiety than others.

**Table 2:** Factors associated with anxiety using log-binomial regression.

| Variables | Unadjusted model |  | Adjusted model |  |
| --- | --- | --- | --- | --- |
|  | Prevalence ratio [95% CI] | p-value | Prevalence ratio [95% CI] | p-value |
| **Age of the respondent** |  |  |  |  |
| 20-24 | 1[0.83,1.20] | 0.002 | 1.01[0.84,1.22] | 0.887 |
| 25-29 | 1.32[1.10,1.57] | <0.001 | 1.32[1.11,1.58] | 0.002 |
| 30-34 | 1.62[1.36,1.91] | <0.001 | 1.55[1.30,1.85] | <0.001 |
| 35-39 | 1.82[1.53,2.16] | <0.001 | 1.69[1.41,2.02] | <0.001 |
| 40-44 | 1.82[1.54,2.15] | <0.001 | 1.65[1.39,1.98] | <0.001 |
| 45-49 | 2.01[1.69,2.38] | <0.001 | 1.77[1.47,2.13] | <0.001 |
| 15-19(RC) | 1 |  | 1 |  |
| **Division** |  |  |  |  |
| Barishal | 1.11[0.89,1.38] | 0.362 | 1.14[0.92,1.42] | 0.217 |
| Chattogram | 1.25[1.02,1.55] | 0.035 | 1.33[1.08,1.64] | 0.007 |
| Dhaka | 0.97[0.80,1.19] | 0.796 | 1.03[0.85,1.25] | 0.774 |
| Khulna | 1.18[0.97,1.44] | 0.106 | 1.22[1.00,1.48] | 0.046 |
| Rajshahi | 1.04[0.84,1.30] | 0.702 | 1.03[0.84,1.27] | 0.766 |
| Rangpur | 1.5[1.23,1.84] | <0.001 | 1.57 [1.30, 1.91] | <0.001 |
| Sylhet | 1.19[0.94,1.51] | 0.157 | 1.2[0.95,1.52] | 0.118 |
| Mymensingh (RC) | 1 |  | 1 |  |
| **Place of residence** |  |  |  |  |
| Rural | 1.12[1.00,1.26] | 0.05 | 0.97[0.87,1.09] | 0.62 |
| Urban | 1 |  | 1 |  |
| **Education of the respondent** |  |  |  |  |
| No education | 2.06[1.80,2.37] | <0.001 | 1.37[1.16,1.62] | <0.001 |
| Primary | 1.72[1.51,1.96] | <0.001 | 1.28[1.10,1.50] | 0.002 |
| Secondary | 1.4[1.23,1.58] | <0.001 | 1.23[1.08,1.41] | 0.003 |
| Higher (RC) | 1 |  | 1 |  |
| **Wealth** |  |  |  |  |
| Poorest | 1.39[1.23,1.58] | <0.001 | 1.21[1.04,1.41] | 0.012 |
| Poorer | 1.35[1.19,1.53] | <0.001 | 1.21[1.05,1.38] | 0.006 |
| Middle | 1.2[1.06,1.34] | 0.003 | 1.09[0.96,1.25] | 0.183 |
| Richer | 1.18[1.05,1.33] | 0.006 | 1.11[0.99,1.25] | 0.087 |
| Richest (RC) | 1 |  | 1 |  |
| **Working status** |  |  |  |  |
| Yes | 1.09[1.00,1.18] | 0.049 | 0.96[0.89,1.04] | 0.362 |
| No (RC) | 1 |  | 1 |  |
| **Has an account in a bank or other financial institution** |  |  |  |  |
| Yes | 1.13[1.03,1.24] | 0.013 | 1.25[1.14,1.38] | <0.001 |
| No (RC) | 1 |  | 1 |  |
| **Participation in decision-making** |  |  |  |  |
| Yes | 1.03[0.96,1.10] | 0.76 | 1.00[0.93,1.07] | 0.953 |
| No (RC) | 1 |  | 1 |  |
| **Attitude toward wife beating** |  |  |  |  |
| Positive | 1.34 [1.22,1.47] | <0.001 | 1.28[1.17,1.40] | <0.001 |
| Negative (RC) | 1 |  | 1 |  |
| **Husband's level of education** |  |  |  |  |
| No education | 1.79 [1.58,2.02] | <0.001 | 1.23[1.05,1.44] | 0.009 |
| Primary | 1.43 [1.28,1.61] | <0.001 | 1.13[0.98,1.30] | 0.086 |
| Secondary | 1.27 [1.14,1.42] | <0.001 | 1.10[0.98,1.24] | 0.112 |
| Higher (RC) | 1 |  | 1 |  |
| **Religion** |  |  |  |  |
| Muslim | 1.29 [1.11,1.50] | 0.001 | 1.40[1.20,1.64] | <0.001 |
| Others (RC) | 1 |  | 1 |  |
| **Exposed to any mass media** |  |  |  |  |
| Yes | 0.9[0.83,0.98] | 0.012 | 1.05[0.97,1.13] | 0.257 |
| No (RC) | 1 |  | 1 |  |
| **Ever used internet** |  |  |  |  |
| No | 1.34[1.22,1.47] | <0.001 | 1.09[0.98,1.22] | 0.097 |
| Yes (RC) | 1 |  | 1 |  |
| RC= Reference category |  |  |  |  |

**Table 3** presents factors associated with depression using log-binomial regression. Women aged 45-19 (PR= 1.73, p=0.004) had a significantly higher prevalence of symptoms of depression. The prevalence of symptoms of depression was significantly highest in women living in Rangpur (PR= 2.14, p=<0.001). Respondents of the poorest wealth quintile had significantly the highest prevalence of depression (PR=1.36, p=0.05). Furthermore, having an account in a bank or other financial institution (PR= 1.60, p=<0.001) caused a significantly higher prevalence of depression. Finally, women who had a positive attitude towards wife beating (PR= 1.47, p=<0.001), who are Muslim (PR= 1.48, p=<0.001), and who had access to mass media (PR=1.20, p=0.016) had a higher prevalence of depression.

**Table 3:** Factors associated with depression using log-binomial regression.

| Variables | Unadjusted model |  | Adjusted model |  |
| --- | --- | --- | --- | --- |
|  | Prevalence ratio [95% CI] | p-value | Prevalence ratio [95% CI] | p-value |
| **Age of the respondent** |  |  |  |  |
| 20-24 | 0.88[0.64,1.23] | 0.459 | 0.92[0.66,1.29] | 0.630 |
| 25-29 | 1.27[.92,1.75] | 0.151 | 1.28[0.91,1.79] | 0.153 |
| 30-34 | 1.42[1.03,1.95] | 0.031 | 1.36[0.98,1.89] | 0.068 |
| 35-39 | 1.49[1.09,2.04] | 0.012 | 1.39[0.99,1.94] | 0.051 |
| 40-44 | 1.47[1.06,2.04] | 0.020 | 1.33[0.93,1.89] | 0.119 |
| 45-49 | 1.99[1.43,2.77] | <0.001 | 1.73[1.19,2.49] | 0.004 |
| 15-19(RC) | 1 |  | 1 |  |
| **Division** |  |  |  |  |
| Barishal | 1.25[0.87,1.81] | 0.232 | 1.31[0.92,1.89] | 0.140 |
| Chattogram | 1.11[0.72,1.72] | 0.628 | 1.20[0.78,1.83] | 0.409 |
| Dhaka | 1.11[0.76,1.60] | 0.591 | 1.15[0.80,1.67] | 0.443 |
| Khulna | 1.74[1.24,2.43] | 0.001 | 1.76[1.26,2.47] | 0.001 |
| Rajshahi | 0.99[0.70,1.42] | 0.978 | 0.97[0.68,1.39] | 0.883 |
| Rangpur | 2.02[1.42,2.88] | <0.001 | 2.14[1.50, 3.04] | <0.001 |
| Sylhet | 1.72[1.14,2.59] | 0.010 | 1.78[1.17,2.68] | 0.006 |
| Mymensingh (RC) | 1 |  | 1 |  |
| **Place of residence** |  |  |  |  |
| Rural | 1.17[0.93,1.46] | 0.183 | 0.98[0.77,1.25] | 0.898 |
| Urban | 1 |  | 1 |  |
| **Education of the respondent** |  |  |  |  |
| No education | 2.15[1.62,2.86] | <0.001 | 1.39[0.96,2.00] | 0.083 |
| Primary | 1.61[1.22,2.11] | 0.001 | 1.16[0.83,1.62] | 0.394 |
| Secondary | 1.49[1.17,1.89] | 0.001 | 1.25[0.95,1.66] | 0.111 |
| Higher (RC) | 1 |  | 1 |  |
| **Wealth** |  |  |  |  |
| Poorest | 1.42[1.10,1.84] | 0.007 | 1.36[1.00,1.85] | 0.050 |
| Poorer | 1.29[0.98,1.70] | 0.067 | 1.24[0.90,1.70] | 0.191 |
| Middle | 1.36[1.05,1.77] | 0.022 | 1.31[0.98,1.76] | 0.067 |
| Richer | 1.15[0.90,1.48] | 0.261 | 1.10[0.85,1.42] | 0.467 |
| Richest (RC) | 1 |  | 1 |  |
| **Working status** |  |  |  |  |
| Yes | 1.05[0.89,1.25] | 0.543 | 0.93[0.79,1.09] | 0.368 |
| No (RC) | 1 |  | 1 |  |
| **Has an account in a bank or other financial institution** |  |  |  |  |
| Yes | 1.35[1.10,1.66] | 0.004 | 1.60[1.30,1.97] | <0.001 |
| No (RC) | 1 |  | 1 |  |
| **Participation in decision-making** |  |  |  |  |
| Yes | 0.90[0.77,1.05] | 0.172 | 0.90[0.77,1.05] | 0.162 |
| No (RC) | 1 |  | 1 |  |
| **Attitude toward wife beating** |  |  |  |  |
| Positive | 1.57 [1.31,1.87] | <0.001 | 1.47[1.24,1.76] | <0.001 |
| Negative (RC) | 1 |  | 1 |  |
| **Husband's level of education** |  |  |  |  |
| No education | 1.88 [1.47,2.41] | <0.001 | 1.40[1.02,1.93] | 0.040 |
| Primary | 1.49 [1.17,1.90] | 0.001 | 1.24[0.92,1.67] | 0.156 |
| Secondary | 1.38 [1.10,1.72] | 0.005 | 1.21[0.93,1.57] | 0.147 |
| Higher (RC) | 1 |  | 1 |  |
| **Religion** |  |  |  |  |
| Muslim | 1.34 [0.99,1.81] | 0.059 | 1.48[1.11,1.98] | <0.001 |
| Others (RC) | 1 |  | 1 |  |
| **Exposed to any mass media** |  |  |  |  |
| Yes | 1.02[0.89,1.18] | 0.795 | 1.20[0.04,1.40] | 0.016 |
| No (RC) | 1 |  | 1 |  |
| **Ever used internet** |  |  |  |  |
| No | 1.36[1.12,1.66] | 0.002 | 1.16[0.95,1.42] | 0.157 |
| Yes (RC) | 1 |  | 1 |  |
| RC= Reference category |  |  |  |  |

## Discussion

This study focused on the prevalence and factors associated with anxiety and depression among reproductive-aged women (15-49) in Bangladesh. The findings reported that 19.5% of respondents had symptoms of anxiety, and 4.9% had symptoms of depression. Furthermore, the findings of our study suggested that age, place of residence, education of the respondent, wealth status, attitude towards domestic violence, spouse’s education, and religion were significant determinants of anxiety among women. Age, place of residence, financial status, attitude towards domestic violence, spousal education, religion, and exposure to mass media were found to have significant associations with depression.

We found that women aged 45-49 had the highest level of anxiety and depression. Studies around the world also found higher levels of anxiety and depression in middle-aged women.^38,39^ Studies in China, Pakistan, and Malaysia found increasing age as a significant determinant of anxiety and depression among women.^40–42^ While increased age can cause poor mental health, many studies found the reverse relationship of the effect of poor mental health on accelerated aging.^43^ Wealth status was found to be significantly associated with anxiety and depression among women in our study. Although some studies found a significant relationship between economic status and anxiety and depression among women, some studies didn’t find any relationship between these two.^44–46^ Poverty was proved to be one of the most consistent predictors of depression among women for its considerable impact on sources of social support for women.^47,48^

Though the educational level of the respondents was significantly associated with symptoms of anxiety, our study didn’t find any significant association between depression and the educational level of respondents. On the contrary, the lower level of education of the spouse of the respondent was an essential predictor of both anxiety and depression among women. While one study in Pakistan found a significant relationship between education and both depression and anxiety, A study in Malaysia only found a significant relationship between depression.^41,42^ Studies concluded that a higher level of education protects women from anxiety and depression by empowering them with stronger predictability values than men.^47,49,50^

Women living in Rangpur had significantly higher levels of anxiety and depression than the rest of the divisions in the country, which the division’s economic situation can explain. Rangpur is the poorest division in the country, and almost half of the population (47.2%) lives in the quintile of poor wealth.^51^ In addition, women who justified wife beating reported significantly higher levels of anxiety and depression than women who didn’t justify wife beating in our study. Studies around the world found that depression, stress-related syndromes, chemical dependency and substance abuse, and suicide are consequences observed in the context of violence in women’s lives.^52^ Studies in Iran, Nigeria, Australia, and Pakistan found domestic violence as a significant risk factor for depression and anxiety among women.^53–56^ Finally, women who have exposure to mass media have a significantly higher prevalence of depression than women who don’t have exposure to mass media. Studies found that exposure to mass media can cause increased depression in women to ideal body image perception, comparisons, and higher expectations, which is predominantly true for adolescent and young women.^57,58^

### Strength and Limitations

This study has compelling strengths as it was the first to explore the prevalence and associated factors associated with mental health using nationally representative data. We analyzed the nationally representative data, which might increase the acceptability and generalization to another setting of similar socioeconomic status. We used log-binomial regression analysis for our study. Log-binomial provides a prevalence ratio that is better for communication than logistic regression.^59^ This study has some crucial limitations, too. Self-reported data is sometimes vulnerable to recall bias and social desirability bias. Moreover, using data from a cross-sectional research design was less than ideal for determining causality. Therefore, a qualitative study and prospective cohort study would bring the best results and explore the factors associated with anxiety and depression among reproductive-aged women in Bangladesh.

## Conclusion

Women of reproductive age face many crucial life events, such as the start of adolescence, childbirth, postpartum, and premenopausal periods, which make them vulnerable to mental health disorders like anxiety and depression.^22,39,46,58^ In this context, our study aims to explore the prevalence and factors associated with anxiety and depression among reproductive-aged women. The high prevalence emphasizes the need for policymakers and healthcare providers to take strategic action towards women’s mental health. Specific focus should be given to women of lower socioeconomic status with limited resource access, explicitly on Rangpur. Moreover, strategic actions should be driven towards women aged 40 or above for their unique needs. A helpline needs to be open for seeking mental health advice. The issues of mental health are still stigmatized in the country. Specifically, the situation is severe for women. For this reason, free and accessible community service should be provided to women for counseling about their mental health. Women facing domestic violence need to be treated separately. In addition, awareness programs should be directed at the whole community to decrease the stigmatization and fear of mental health problems in the community and society. Mass media programs should be designed to meet the needs of women, and harmful practices and cultures like body image perception, comparisons, and higher expectations should not be promoted.

## Data Availability

Data is publicly available at DHS website (https://www.dhsprogram.com/data/available-datasets.cfm), and you can access to data following some procedure.

